# Cognitive AI-Assisted Primary Care Health Delivery: A Pilot Study in Bangladesh

**DOI:** 10.64898/2026.04.03.26349253

**Authors:** Raeed Kabir, Michael Williams, Nashif Rayhan

## Abstract

Research has documented persistent physician workforce shortages globally, with projected shortfalls threatening primary care access in underserved populations. Existing AI applications in healthcare have largely focused on predictive risk-scoring tools that generate probability estimates but do not reduce the time a physician spends completing a patient encounter. A 2026 working paper further demonstrated that large language models lack the metacognitive capacity necessary for reliable medical reasoning, i.e., being able to ask appropriate questions in the absence of information to collect patient history and update differential diagnoses. This paper reports on a 2025 pilot deployment of ClinicalAssist in Bangladesh that tested a fundamentally different model: an AI system designed to replicate every step of the clinical workflow. Across 239 unique patients, 277 encounters, and 287 diagnostic opportunities, the system achieved an overall diagnostic accuracy of 94.7%, with chronic disease accuracy of 98.0% and acute care accuracy of 88.9%. These results suggest that cognitive AI has the potential to be a powerful clinical force multiplier if properly integrated in workflow.

## 1 Introduction

Healthcare systems in low- and middle-income countries face a structural paradox: demand for primary care continues to rise while the supply of trained physicians stagnates or contracts. Research has documented persistent workforce shortages ([1], [2], [3]): academic analyses project a physician deficit of up to 86,000 in the United States by 2036, while the Physicians Foundation Scorecard (2024) documents capacity limitations and a workforce exodus from primary care more broadly. ClinicalAssist, a U.S. based company provided the AI software studied in this paper. The AI will henceforth be referred to as “ClinicalAssist,” after its parent company, though it is sometimes called Doctor AI, its original name. Doctor AI operates by predicting the next best question a seasoned clinician would ask, balancing the patient’s clinical presentation with diagnostic cost-effectiveness. By mastering this predictive sequence, the system effectively replicates a physician’s professional cognitive process while simultaneously identifying the most likely diagnosis, treatment strategies, and associated encounter notes. The software was piloted in Bangladesh, in particular, due to its landscape of acute structural deficiency in healthcare. Bangladesh has approximately five physicians per 10,000 population nationally, but this figure masks a severe rural-urban disparity: in rural areas the ratio falls to roughly 21 doctors per 100,000 people, compared to 249 per 100,000 in urban centers, meaning rural Bangladeshis face a physician availability gap of more than tenfold [4]. More than 75% of the rural population turn instead to non-physician providers as their first point of care, a category that includes approximately 12 unqualified village doctors per 10,000 population [4].These informal providers operate alongside an equally unregulated pharmaceutical market: research conducted in rural Bangladesh identified 301 antibiotic dispensers in a single catchment area, of whom 52% operated without legal authorization, and a separate cross-sectional survey found that 56.6% of antibiotics in Bangladesh are purchased without a physician’s prescription, often dispensed directly by drug-shop operators to customers who frequently cannot distinguish between a registered physician and an unlicensed vendor ([5], [6]). The practical consequence is a primary care environment in which the boundary between commerce and medicine has effectively dissolved, and in which the absence of a qualified physician is not a barrier to receiving treatment.

Long waiting times and access barriers compound the problem. Research has shown that waiting time functions as a sentinel indicator for health services under strain [7], with delays in care access disproportionately affecting rural and low-income populations—precisely the demographics that the Bangladesh pilot serves. The policy response has increasingly focused on AI and machine learning applied to clinical data. The dominant paradigm is predictive: tools that generate probability estimates of mortality risk, readmission likelihood, or disease progression. A system predicting sepsis mortality risk from pneumonia or UTI can be useful for population-level stratification, but a probability score does not reduce the time a physician spends on a patient encounter. The physician still must take a history, arrive at a diagnosis, construct a treatment plan, and produce documentation. Predictive AI assists with none of these tasks.

A major external validation study found the Epic Sepsis Model failed to identify about two-thirds of actual sepsis patients. There were frequent false positives, which can desensitize clinicians and contribute to alert fatigue. Researchers found the model had poor discrimination (inability to separate high-risk from low-risk patients) and poor calibration (predicted risks didn’t match actual outcomes) [8]. A January 2025 study in *Nature Communications* demonstrated that large language models lack essential metacognition for reliable medical reasoning: they cannot reliably identify the limits of their own knowledge, a property essential for safe clinical deployment [9]. This finding reinforces the distinction between AI systems that operate as consultative overlays and those structurally integrated into the physician’s workflow under physician supervision. As Dr. Azad Kabir, developer of ClinicalAssist, explained in an interview cited by *Wired* [10]: “Most AI models aren’t built for medicine. They lack the ability to build a case scenario from one symptom, question themselves, analyze alternatives, auto-correct errors or erroneous data, and assess risk dynamically.”

ClinicalAssist was developed to address this gap. Rather than predicting a single outcome, it replicates every step of a clinical visit: strategic history-taking to generate and narrow a differential diagnosis, structured reasoning toward a confirmed diagnosis, evidence-based treatment planning, and automated generation of an encounter note without needing involvement of a physician. When AI matches the physician’s workflow from start to finish, encounter time drops substantially and patient throughput rises. The efficiency gain that transforms healthcare is not measured in risk scores, but it is measured in physician time. In a recent study, the capability of the triage capacity of the ChatGPT Health LLM to triage emergency clinical scenarios was evaluated, and this showed that among well-studied gold-standard emergency scenarios, the system under-triaged 52% of cases, directing patients with diabetic ketoacidosis or impending respiratory failure to 24–48 hour follow-up evaluation rather than the emergency department, while correctly triaging classical emergencies such as stroke and anaphylaxis [11].

Popular technologies exist that use AI to scribe conversations that require physicians to be present in the first place. These are stated to be force multipliers, but as [12] argue, a system that cannot operate independently at any stage of the clinician workflow cannot function as a force multiplier, and the global shortage of primary care can only be addressed by tools that reduce the need for clinician presence rather than merely augmenting it. ClinicalAssist operates without requiring clinician presence during data collection, and it is this independence that constitutes its primary structural advantage over ambient scribe technology or risk prediction systems.

This paper reports on the 2025 Bangladesh pilot of ClinicalAssist: the first systematic field deployment of the system in a live clinical environment, spanning January through December 2025 across two sites in Comilla District. The pilot enrolled 239 unique patients across 277 clinical encounters; some encounters included multiple diagnoses, though single-diagnosis presentations were the norm.

## 2 The ClinicalAssist Workflow

ClinicalAssist is organized around four discrete phases of the physician encounter, each supported by targeted AI functionality. The proportional time allocations reflect observed patterns in primary care practice and represent the loci of efficiency gain.

### Step 1: Patient History

The system employs strategic questioning logic to identify the next best question at each turn, efficiently narrowing the differential diagnosis. This is perhaps the largest gap between existing predictive models and the ClinicalAssist model that does not rely upon training data. At its current level, ClinicalAssist has more than 15,000 diagnoses linked with more than 2,200 signs, symptoms, and laboratory and radiological findings. In addition, it provides complete decision support for the most common 400 medical conditions. These 400 medical conditions are chosen from a list of commonly presented conditions in urgent care centers, urgent care and primary care clinics, and hospitalized patients. Within approximately two minutes of symptom input, it generates a short list of candidate diagnoses and links clinical findings to each candidate. This mirrors the information-gathering behavior of an experienced clinician without needing the presence of a physician.^1^ Notably, ClinicalAssist’s model was unchanged between U.S. and Bangla contexts, as it is not trained on disease prevalence data but using weights determined by a panel of expert physicians that can connect symptomatology to disease states.

### Step 2: Diagnosis Finalization

The system revisits clinical findings to confirm or rule out each differential candidate, attempting to exclude the most probable diagnosis before accepting it. The output is a ranked, evidence-linked diagnosis with supporting clinical rationale, which is designed to reduce anchoring bias.^2^

### Step 3: Treatment Plan

The system generates a treatment plan from the confirmed diagnosis, drawn from up-to-date evidence-based protocols. Physicians review and select from structured options; the system does not prescribe autonomously. This reduces the cognitive load of treatment selection while preserving full physician authority over clinical decisions.

### Step 4: Documentation

Medical documentation, the construction of comprehensive encounter notes for medico-legal purposes, accounts for approximately half of total physician encounter time. ClinicalAssist automates encounter note generation from the structured data collected in Steps 1–3, producing a complete clinical record without separate physician dictation or transcription. This is the single largest source of time savings in the workflow.

### Triage Framework

After determining the diagnosis, and in conjunction with basic vitals data, ClinicalAssist generates a severity score that bins individuals into various triage tiers. Triage Type 1 designates emergent cases with immediate mortality risk requiring hospitalization without delay. Triage Type 2 designates cases in which the patient waits for physician validation— analogous to standard emergency triage performed by nursing staff. Triage Type 3 cases are the lowest priority. Predicting risk of death from sepsis, pneumonia, or UTI is possible with sufficient data input but yields only a single probabilistic outcome; the triage framework operationalizes this risk stratification into a clinically actionable, workflow-integrated form.

## 3 Pilot Study Design and Context

The 2025 Bangladesh pilot was a prospective observational deployment study with two objectives: testing the technical viability of AI-powered telemedicine in resource-limited settings, and generating initial clinical performance data against physician-validated ground truth. The pilot ran continuously from January through December 2025 across two phases.

### Phase 1: Barura, Comilla (January–April 2025)

The first phase established a free satellite clinic in Barura, a rural community in Comilla District, structured as a walk-in village clinic analogous to an urgent care facility. The station was equipped with dedicated internet connectivity, a full suite of vital measurement instruments, an electronic stethoscope with remote auscultation capability, and electrocardiography (EKG) analytics. This infrastructure allowed the physician-AI system to function with the diagnostic capacity of a well-resourced primary care clinic despite its rural location. Patient volume during Phase 1 was lower than projected. Both the Barura site and the subsequent Comilla site were newly established satellite facilities without prior community presence or reputational history. Community trust in clinical institutions in rural Bangladesh builds incrementally and is contingent on established provider relationships—a structural constraint for any newly opened facility, and not one specific to AI-assisted care.

### Phase 2: BSCIC Industrial Area, Comilla (April–December 2025)

In April 2025, the pilot transitioned to the Bangladesh Small and Cottage Industries Corporation (BSCIC) industrial area in Comilla, serving industrial workers and their dependents— a semi-urban population with distinct epidemiological characteristics relative to the rural Barura catchment.

The first half of Phase 2 focused on acute care for patients with new, urgent symptoms. The second half shifted deliberately toward chronic disease management to rigorously stress-test the ClinicalAssist model under the more complex, longitudinal conditions of ongoing NCD care. To address local reluctance among chronically ill, often asymptomatic patients to pay for regular follow-up, chronic care was provided at no cost under certified physician supervision. Acute care was offered for a minimal fee. Total unique patient volume reached 239 across both phases and the full year.

## 4 Patient Population and Data

Over twelve months, 239 unique patients were treated across 277 clinical encounters. Some patients presented on multiple occasions; some encounters involved more than one diagnosis, though single-diagnosis presentations were the norm. Thus, there are 287 diagnostic opportunities, as each diagnosis is an independent exercise for the ClinicalAssist AI model. The patient history data was strictly collected by ClinicalAssist, but the clinical encounter data was collected by the supervising physician, Dr. Nashif Rayhan (a Bangladeshi native primary care physician)^3^. Data was entered into an Excel sheet, which contained a daily log recording patient identifiers, presenting symptoms, the AI system’s diagnostic and treatment output, and a binary accuracy assessment of whether the system’s diagnosis or treatment plan was validated by the clinician’s judgment. All encounters were classified as acute or chronic based on the nature of the presenting condition. For chronic encounters, the relevant diagnosis is sometimes known at presentation; the physician’s task is to assess whether the condition is stable, improving, or deteriorating. AI assistance in structured history collection and documentation therefore remains valuable even when diagnostic novelty is not the primary clinical challenge.

The pilot period also served as a live debugging environment. A software issue in the first half of the pilot prevented some treatment plans from being saved to the electronic health database stored by the software; this was corrected in mid-June 2025, and all treatment plan data reflect the post-correction period.

## 5 Results

### 5.1 Diagnostic Accuracy

|  |  |  |
| --- | --- | --- |
| **98.0%** | **88.9%** | **15** |
| **Chronic Accuracy** | **Acute Accuracy** | **Excluded Records** |
| 200 of 204 | 72 of 81 | Unmatched, wrong diagnosis,<br>wrong treatment, 2 not<br>labeled as chronic/acute. |
| **272** | **94.7%** | **239** |
| **Successful Diagnoses** | **Overall Accuracy** | **Unique Patients** |
| After 15 exclusions | 272 of 287 correct | Across 277 encounters |

Over the sample period, 239 unique patients came to the clinic to be seen (by the AI). This resulted in 277 encounters, as there were some returners, and each patient encounter could involve multiple diseases and opportunities for ClinicalAssist to work up the patient. Thus, there were 287 total records. 285 were classified as acute or chronic. 15 of 287 were excluded on the basis of physician-flagged diagnostic errors, cases where the AI output could not be matched to a valid clinical entity, represented a documented wrong diagnosis, or involved a missing diagnosis entirely. The remaining 272 encounters formed the analysis sample, with the accuracy denominator set at 285 to reflect all classified records. Notice the discrepancy between the 285 and 287 total diagnosis figures. The two diagnoses that had missing labels for acute vs chronic were treated as “incorrect diagnoses” to be conservative about accuracy and to align the number of successful diagnoses (272) with the chronic (200) and acute (72) breakdown. Note that some diagnoses, like anxiety and depression, are not always one or the other. These are noted as chronic unless proven as acute.

Across all 285 classified (as acute or chronic) encounters, ClinicalAssist achieved an overall diagnostic accuracy of 94.7% (272 correct) [91.5%, 97.0%]^4^. Accuracy differed meaningfully by encounter type. Among 204 chronic disease encounters, 200 were correctly diagnosed, yielding a chronic accuracy rate of 98.0% [95.1%, 99.5%]. Among 81 acute care encounters, 72 were correctly diagnosed, yielding an acute accuracy rate of 88.9% [80.0%, 94.8%].

The overall error rate was 5.3%. Data on the correct diagnosis were not always collected if the physician indicated a discrepancy between their clinical assessment, so we are only able to calculate “false positives” at a disease level. For each diagnosis X, we define the condition-level false discovery rate as the proportion of AI-positive claims for X that were not validated by the supervising physician. The false discovery rate for the five highest-volume diagnoses (hypertension (n=58), diabetes mellitus (n=50), asthma (n=17), diabetic foot ulcer (n=15), and osteoarthritis (n=14)) were each 0.0%. The highest false discovery rate among diagnoses with more than one claim was scabies at 4.2% (1/24).

**Figure 1:**
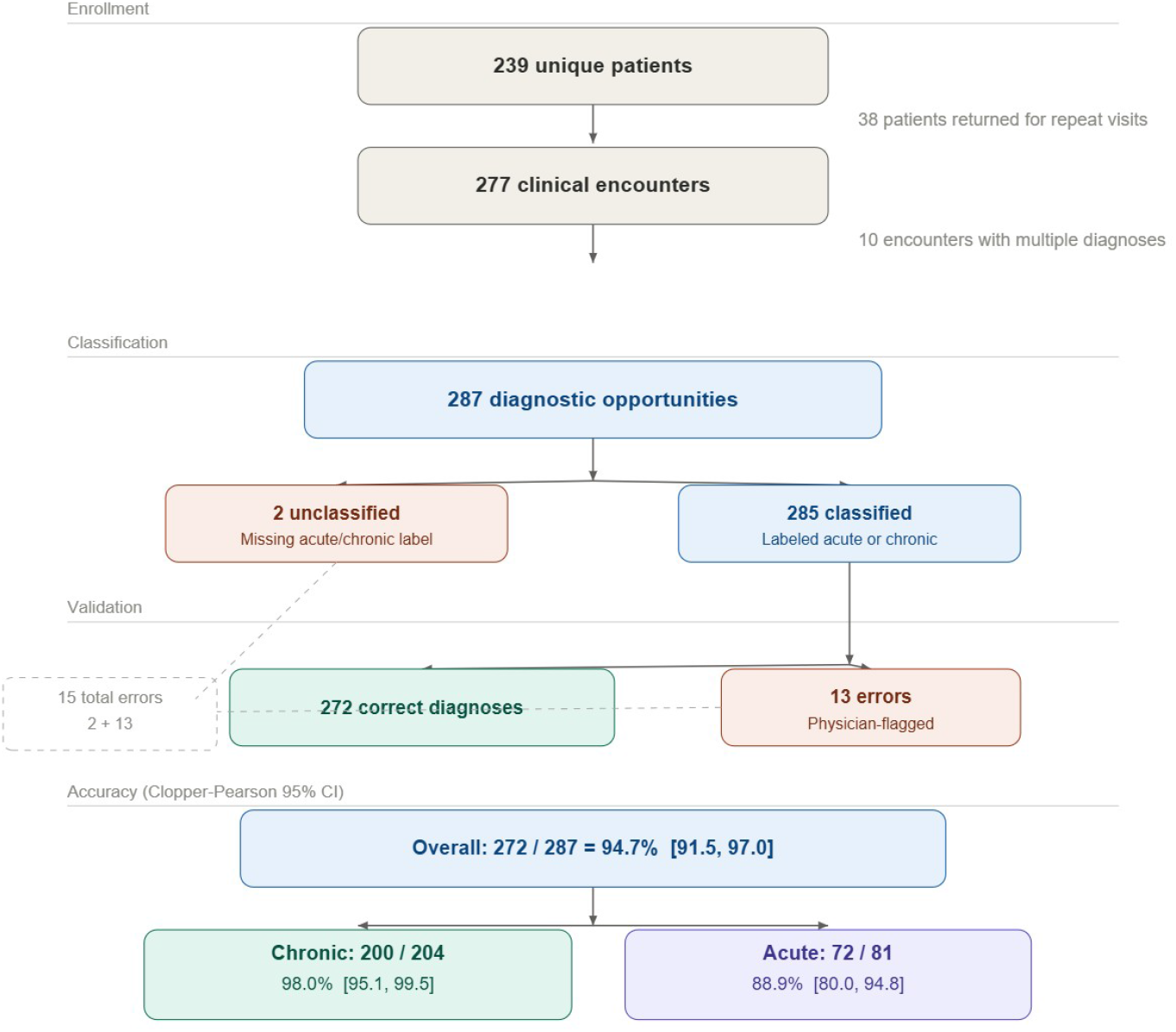
Study sample breakdown.

The differential between chronic and acute accuracy is clinically interpretable. Chronic conditions present with known diagnoses and stable symptom patterns; the AI’s task is to support structured history collection and stability assessment, not generate a novel diagnosis from an undifferentiated symptom cluster. Acute presentations involve novel symptom patterns, overlapping differentials, and broader diagnostic uncertainty. An 88.9% first-year accuracy rate in acute care is a meaningful result for a pilot deployment.

### 5.2 Chronic Disease Encounters

After aggregating related diagnoses—consolidating hypertension and hypertension follow-up visits, collapsing variant spellings of diabetes mellitus, merging diabetic foot ulcer and follow-up records, and combining scabies with infected scabies—the chronic encounter sample comprised 200 visits across 25 unique diagnostic categories.

**Table 1:** Chronic encounter distribution by aggregated diagnosis. Hypertension and diabetes mellitus together accounted for 53.5% of all chronic visits. The top five diagnoses covered 76.5% of chronic encounters.

| Diagnosis (aggregated) | Visits | % of Chronic |
| --- | --- | --- |
| Hypertension (incl. follow-up) | 58 | 29.0% |
| Diabetes mellitus (all variants) | 49 | 24.5% |
| Asthma | 17 | 8.5% |
| Diabetic foot ulcer (incl. follow-up) | 15 | 7.5% |
| Osteoarthritis | 14 | 7.0% |
| Ankylosing spondylitis | 8 | 4.0% |
| Sciatica | 6 | 3.0% |
| Anxiety | 5 | 2.5% |
| Hypothyroidism | 5 | 2.5% |
| Peptic ulcer | 4 | 2.0% |
| Other (15 diagnoses, $\leq 2$ visits each) | 19 | 9.5% |

The five most common chronic diagnoses, hypertension, diabetes mellitus, asthma, diabetic foot ulcer, and osteoarthritis, accounted for 76.5% of all chronic encounters. This concentration reflects the epidemiological burden of non-communicable diseases in the pilot population, consistent with documented patterns in South Asian primary care. Hypertension alone represented 29.0% of chronic visits; combined with diabetes mellitus, the two conditions accounted for 53.5% of all chronic encounters.

### 5.3 Acute Care Encounters

The acute encounter sample comprised 72 visits across 25 unique diagnostic categories after aggregation of scabies variants, urinary tract infection entries, and tinea subtypes.

Scabies was the dominant acute presentation, accounting for 30.6% of all acute encounters— a proportion reflecting the BSCIC industrial setting, where close working and living quarters facilitate transmission of skin infestations. Respiratory conditions—pneumonia, influenza, and viral syndrome combined—accounted for an additional 22.2% of acute visits. The top five acute diagnoses collectively covered 59.7% of encounters, with the remainder distributed across 20 additional categories including dengue fever, acute pancreatitis, cerebrovascular accident, and diverticulitis.

**Table 2:** Acute encounter distribution by aggregated diagnosis. Scabies accounted for 30.6% of acute visits, reflecting the epidemiological context of an industrial worker population. The top five diagnoses covered 59.7% of acute encounters.

| Diagnosis (aggregated) | Visits | % of Acute |
| --- | --- | --- |
| Scabies (incl. infected) | 22 | 30.6% |
| Pneumonia | 7 | 9.7% |
| Urinary tract infection (all variants) | 5 | 6.9% |
| Influenza | 5 | 6.9% |
| Viral syndrome | 4 | 5.6% |
| Gastroenteritis | 3 | 4.2% |
| Strep throat | 3 | 4.2% |
| Tinea (all subtypes) | 3 | 4.2% |
| Otitis externa | 2 | 2.8% |
| Dengue fever | 2 | 2.8% |
| Other (15 diagnoses, 1 visit each) | 15 | 20.8% |

## 6 AI Generated Treatment Plans

Treatment plan performance was not a primary focus of the 2025 Bangladesh pilot, as the study was principally designed to evaluate diagnostic accuracy against a physician reference standard. Nevertheless, the available treatment data are encouraging. Of the 287 recorded encounter rows, 221 (77.0%) had a confirmed saved treatment plan. This figure understates true treatment performance: a software bug in the first half of the pilot prevented treatment plans from being logged, a limitation corrected in mid-June 2025, meaning a substantial portion of the 64 unsaved encounters reflect a data capture failure rather than a clinical one. Among the encounters where the diagnosis was correct and a comment was recorded, 216/221 cases, i.e., an accuracy bounded between [94.8%, 99.3%], had correct treatment plans. That is, 216/221 cases were not reported as having a treatment plan with which the physician disagreed — specifically, the system could not produce an appropriate treatment plan for burn care, rheumatoid arthritis, erectile dysfunction and premature ejaculation, impetigo, and scabies in one instance each. This suggests that treatment failures were concentrated in a small set of lower-frequency or non-standard conditions outside the system’s primary design envelope. For the high-volume conditions that dominated the pilot — hypertension, diabetes mellitus, asthma, and diabetic foot ulcer, which together accounted for over half of all encounters — the treatment plan accuracy mirrors the near-perfect diagnostic accuracy reported above. Formal treatment plan evaluation, including structured comparison against evidence-based protocols, is planned as a primary outcome in later work.

## 7 Discussion

The results of the 2025 Bangladesh pilot support two primary conclusions. First, ClinicalAssist demonstrated clinical accuracy sufficient for real-world deployment as a physician-support tool, achieving 94.7% overall diagnostic accuracy in a live clinical environment with genuine patient populations and active physician supervision. Second, the accuracy differential between chronic (98.0%) and acute (88.9%) presentations is consistent with the structural difference in diagnostic complexity between the two encounter types and provides a useful baseline for targeted system improvement.

The chronic accuracy figure is particularly significant because it was achieved in a context where the AI’s primary value was not diagnostic novelty but clinical workflow support: structured history collection, stability assessment, and automated documentation. That 200 of 204 chronic encounters were correctly handled demonstrates that the system can reliably manage the most common chronic conditions in a South Asian primary care population— hypertension, diabetes, and asthma—which together represent the leading drivers of NCD morbidity in the region. Of the four errors, 1 was coded as an error because of a software bug that prevented adding muscle pain for a depression diagnosis, one patient encounter with an undetermined diagnosis, and two conditions that were diagnosed by the supervising physician but not by the AI: penile ulcer and chronic kidney disease (CKD). To be conservative, any errors regarding the history collection and diagnosis process were counted as diagnostic errors. This is meant to simulate a failure when an assistant with less clinical training (like a community paramedic or community health worker) is using the software to engage patients.

The acute accuracy figure of 88.9%, while lower, reflects a harder diagnostic problem. Acute presentations frequently involve undifferentiated symptom clusters in which a febrile patient could present with influenza, dengue, pneumonia, or viral syndrome. The 9 incorrect cases among 81 acute encounters represent the system’s primary performance frontier; analysis of excluded and incorrect cases reveals that the most common failure modes were diagnostic mismatch in ambiguous presentations and the absence of a code-able diagnosis—not systematic misclassification of well-defined conditions.

The chronic accuracy statistic may also be higher because chronic follow-up appointments are counted as a “successful” diagnoses, despite the patient already knowing the answer. To test the accuracy of de novo chronic encounters, we exclude 35 follow-ups. This retains all 4 incorrect diagnoses and gives a 165/169 = 97.6% accuracy rate. Thus, the accuracy rate is robust to accuracy inflation from follow-ups.

The concentration of encounters in a small number of disease categories, where the top five chronic diagnoses covering 76.5% of chronic visits and the top five acute covering 59.7% of acute visits, has direct implications for optimization. Hypertension, diabetes mellitus, asthma, and scabies together account for a disproportionate share of encounters from just the sample population. This reflects a general phenomenon in disease frequency: a small set of conditions covers the overwhelming majority of clinical encounters. It is in these settings that ClinicalAssist can be the most effective.

The pilot also surfaced important operational lessons. Patient volume was lower than projected at both sites, attributable principally to the absence of established community presence at newly opened satellite clinics. This is a structural challenge for any newly deployed healthcare facility; future deployments should be integrated into existing clinical infrastructure where possible rather than launched as de novo satellite centers. The mid-year software correction restoring treatment plan saving further highlights the importance of robust data infrastructure monitoring during live deployment phases.

The performance demonstrated in this pilot was achieved not by generating a risk score but by actively supporting history-taking, diagnosis, treatment planning, and documentation. The operative measure of clinical efficiency is physician time per encounter, and ClinicalAssist is designed to reduce that time at every step. The 2025 Bangladesh pilot provides the first field evidence for this model—echoing the vision articulated in *Wired* [10] that cognitive AI systems, rather than predictive ones, represent the next frontier of clinical impact.

**Figure 2:**
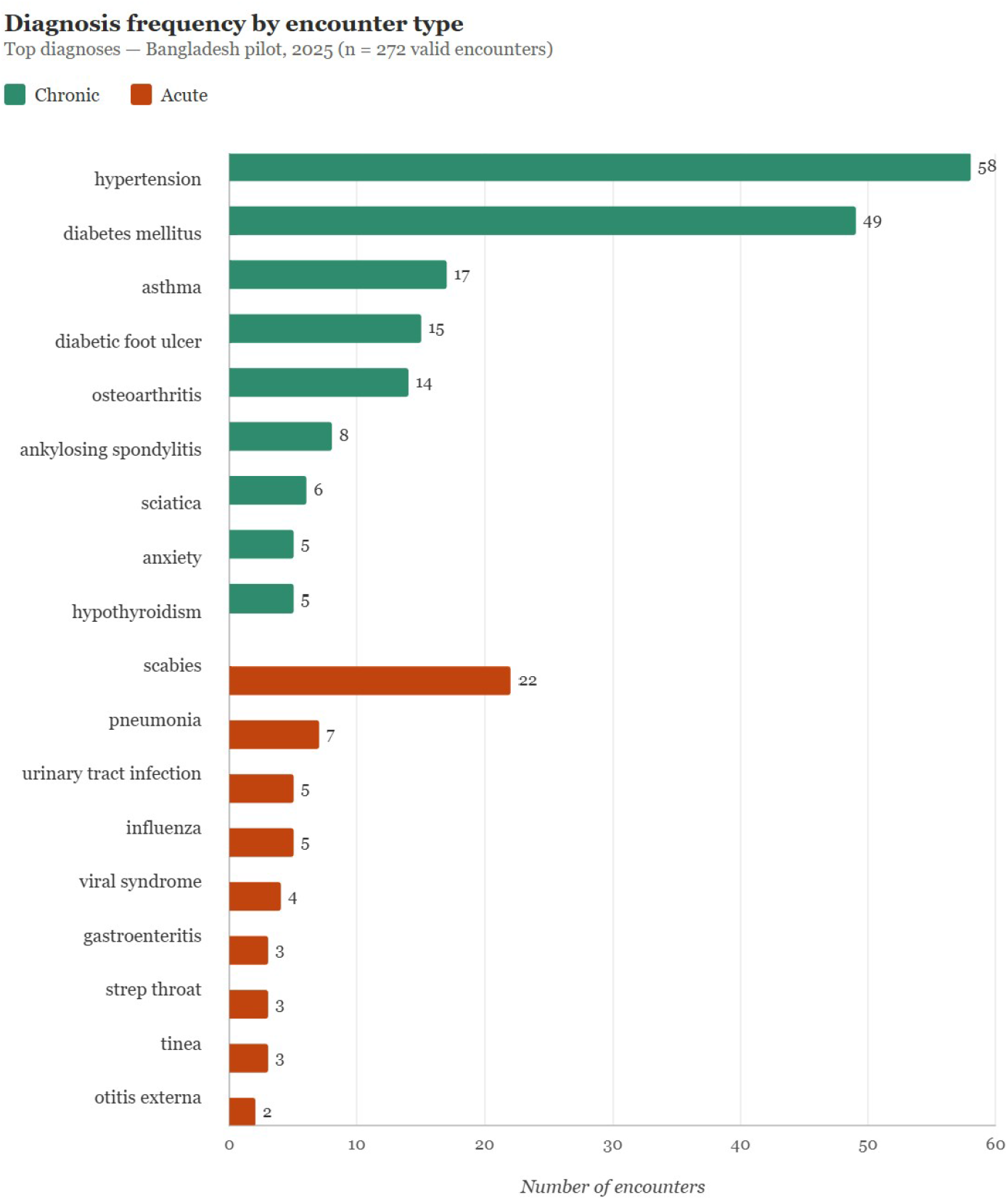
Top diagnoses. Condition names are aggregated across variant spellings and follow-up entries. Remaining diagnoses (*n <* 3) not shown.

## 8 Limitations and Future Directions

Several limitations of the current pilot should be acknowledged. First, the accuracy metric reflects physician validation of the AI’s primary diagnosis—not a comprehensive assessment of the full clinical reasoning chain. This is addressed partially by the accuracy measures on treatment plan. Future evaluations should include structured assessment of the reasoning pathway, not only the diagnostic output. Moreover, there was only one physician, albeit a native Bangladeshi physician, supervising the AI model. Future work should use a panel of expert physicians to validate the model.

Second, the pilot population was drawn from two specific settings that may not be representative of the broader range of South Asian primary care environments. The high prevalence of scabies in the acute sample is likely specific to the BSCIC industrial context; performance across different epidemiological settings should be assessed before generalization.

Third, the pilot was conducted as an observational deployment study without a control condition. A randomized controlled evaluation comparing physician encounters with and without AI assistance on speed, accuracy, and patient outcomes would provide substantially stronger evidence of the system’s impact.

Fourth, the sample of 272 matched diagnoses across 239 unique patients is sufficient for proof-of-concept but underpowered for subgroup analyses or formal statistical inference about condition-specific accuracy rates. Expansion to multiple sites and larger patient volumes is a priority for the next phase.

Future research directions include: a multi-site randomized evaluation of physician encounter time and patient outcomes with and without ClinicalAssist; prospective assessment of documentation quality and completeness; evaluation in additional clinical contexts including emergency triage and specialist referral; and formal economic analysis of the value of encounter time reduction at the system level.

## 9 Conclusion

Project ClinicalAssist represents the first field deployment of ClinicalAssist in a live clinical environment. With 272 matched diagnoses spanning acute and chronic conditions in rural and industrial settings in Bangladesh, the system achieved an overall diagnostic accuracy of 94.7%, with chronic disease accuracy of 98.0% and acute care accuracy of 88.9%. These results—generated under real-world conditions with genuine patient populations and physician supervision—establish that a workflow-integrated AI system, one designed to replicate the full physician encounter rather than overlay a predictive score, can achieve clinical-grade performance as a physician support tool.

The efficiency gain that can transform healthcare in underserved settings is not measured in probability scores. It is measured in physician time per encounter and in the number of patients a physician can see in a single day. ClinicalAssist is designed to reduce the former and increase the latter at every step of the clinical workflow. The 2025 Bangladesh pilot demonstrates that this model is operationally viable. The next phase of research will determine how large the efficiency gains are and for whom they matter most.

## Data Availability

De-identified data is available upon reasonable request to the authors.

## Footnotes

1 This is protected by granted patent.

2 This step is also covered by granted patent.

3 Dr. Rayhan was compensated by ClinicalAssist to run the clinic for 1 year and provide the standard of care to any patients who came to run the satellite sites. This paper is a retrospective analysis of the patients seen by Dr. Rayhan, as he was assisted in every patient encounter by AI, from patient history collection to treatment plan and encounter note generation.

4 Confidence intervals use 95% exact binomial (Clopper-Pearson) bounds.

